# How the COVID-19 pandemic has adversely affected the economics of U.S. emergency care

**DOI:** 10.1101/2020.12.12.20248102

**Authors:** Jesse M. Pines, Mark S. Zocchi, Bernard S. Black, Rebecca Kornas, Pablo Celedon, Ali Moghtaderi, Arvind Venkat, For the US Acute Care Solutions Research Group

**Affiliations:** US Acute Care Solutions, Canton, OH; Department of Emergency Medicine, Allegheny Health Network, Pittsburgh, PA; The Heller School for Social Policy and Management, Brandeis University, Waltham, MA; Northwestern University, Pritzker School of Law and Kellogg School of Management, Evanston, IL; Centura Health, Golden, CO; Milken Institute School of Public Health, George Washington University, Washington, DC

**Author notes:** Funding/Support: No funding was secured for this study.

## Abstract

**Objective:** We describe how the coronavirus (COVID-19) pandemic impacted emergency department (ED) economics, acuity, and staffing.

**Methods:** We conducted an observational study of visits during January to September 2020 compared to 2019 in 136 EDs staffed by a national emergency medicine group. We created ratios of three-week moving averages for 2020 visits, acuity, costs divided by 2019 moving averages, by age and ED size. We tabulated reductions in clinician hours and FTEs compared to early 2020 staffing.

**Results:** 2020-2019 ED visit ratios declined in March nadiring mid-April for both adults (to 0.60) and children (to 0.30) and rose thereafter but remained below 2019 levels through September 2020. The ratio of adult RVUs/visit rose to 1.1 for adults and 1.2 for children in the early pandemic, falling to 1.04 and 1.1 through September. The ratio of direct salary expenses in freestanding (FSED) and small EDs declined less dramatically than in medium and large EDs. Clinical revenues in medium and large EDs declined more sharply and recovered slowly but plateaued well below 2019 levels. By September 2020, expenses were still higher than revenues for small EDs, similar for FSEDs, and somewhat higher for medium and large EDs. During the pandemic, physician hours fell 15% and APP hours 27% during COVID-19 translating to 174 lost physician and 193 lost APP FTEs.

**Conclusion:** The COVID-19 pandemic reduced ED visits and increased acuity in the first 7 months of the pandemic, leading to a contraction of the ED workforce, and threatening ED economics, more so in small and FSEDs.

## INTRODUCTION

### Background

In early 2020, the coronavirus disease 2019 (COVID-19) pandemic spread throughout the world. In the United States and other countries, there were reports of substantial numbers of people delaying or avoiding necessary emergency care in the early pandemic.^1^ Avoidance of emergency care was driven by stay-at-home orders, concerns the healthcare system would be overwhelmed by COVID-19 cases, and personal concerns about risks of viral transmission in emergency departments (ED).^2^ Starting mid-March, there was a precipitous decline in U.S. ED visits. Visit rates reached a low point in the second week of April at 58% of 2019 volumes.^3,4^ Since that time, nationwide ED visits have partially recovered. Yet they still remain substantially below 2019 levels. Other potential contributors to lower ED visits include less communicable disease and injury due to public health measures, including social distancing, mask wearing, and school/business closures. In addition, some care may have shifted to other venues, specifically telemedicine.^5^

### Importance

Medical care delivery by emergency physicians and advanced practice providers (APPs) in EDs is primarily reimbursed by payers on a fee-for-service basis for each individual case. Different models exist through practice organizations to pay clinicians, including fixed hourly rates, payments for generating relative value units (RVUs) – a measure of the number of patients and volume of work completed, or a combination of the two models (hourly plus RVU incentive). Therefore, the economics of practice organizations are directly dependent on patient volumes. Therefore, practice organizations staff EDs to meet volume demands, which includes increasing staffing as volume rises and decreasing staffing with volume declines. When volumes fall as with COVID-19 pandemic, this threatens the economic viability of ED practice organizations, as well as the salaries and jobs of ED clinicians, who are paid hourly or by RVU.

### Goals of the investigation

To our knowledge, no studies have quantified the economic impact of COVID-19 pandemic on ED practice organizations or ED clinicians. Here we describe how visits have evolved during the COVID-19 pandemic, how acuity has changed in EDs, and how this has impacted the economics and staffing in EDs.

## METHODS

### Study Design and Settings

We conducted an observational study of visit rates during January-September 2020, compared to the same period during 2019 in 136 EDs who contracted with a national emergency medicine group for ED clinician services. Facilities include small EDs (<30,000 visits/year, n=40), medium EDs (30,000-59,000 visits/year, n = 56), large EDs (≥60,000 visits/year, n=16), and free standing EDs (FSED) (n=24). These facilities are geographically diverse, located in 18 states. Of the 112 hospital-based EDs, 104 (93%) are non-academic community hospitals. This study was determined to not be human subjects’ research by the Institutional Review Board at Allegheny General Hospital.

### Outcomes & Study Data

Outcomes include the visit numbers, visit acuity (average RVUs per visit), clinical revenue, direct salary expenses for clinicians, clinical hours worked, and full-time equivalents (based on a minimum of 108 hours per clinician per month which is required to obtain benefits). We used data drawn from the national emergency medicine group’s data warehouse for EDs that remained open and staffed in both 2019 and 2020 (balanced panel). This included patient-level data, demographics, billing and reimbursement, clinician payments, and hours worked. The group receives data directly from electronic health records at each site. Reported revenue is the expected revenue because as of this study, actual revenue has not been fully collected. Expected revenue is estimated based on the normal timing and execution by the group’s revenue cycle management functions, which historically accurately tracks with actual revenue.

Data are aggregated to the facility-week level for January-September 2019 and 2020 (39 weeks in each year). All reported weeks begin on Sunday and end on Saturday. We drop Jan 1-Jan 4 in 2020 and Jan 1 – 5 in 2019. Data on staffing expenses are only available at the monthly level. Data on expenses are collected at the facility-month level.

### Methods of Measurement and Data Analysis

We created three-week moving averages for each variable (i.e., we report for week t the average of weeks t-2, t-1, and t]) for each ED. We then computed a 2020/2019 ratio by dividing the 2020 moving average by the 2019 moving average for the corresponding 2019 weeks. We winsorized ratios at the 1^st^ and 99^th^ percentiles to reduce outliers. We then plotted means and 95% confidence intervals for the ratios, stratified by patient age or ED size.

We examined means and ratios during specific time periods related to the COVID-19 pandemic. The pre-pandemic time period consisted of weeks 2-11 (January 5 – March 14). With the exception of the last two days of week 11, this time period fell prior to the national emergency declaration of March 13^th^. We then considered the next 6 weeks of the pandemic (March 15 – April 25) as the “early” COVID-19 period when many states imposed stay-at-home orders, closing schools and non-essential businesses. The week 12 moving average (ending March 21) covered March 1-21 and the week 13 moving average covered March 8-28, so both weeks include both pre-pandemic and during-pandemic data. The remainder of the study period (weeks 18-39) make up the final time period of interest, aligning with general reopening and increased social mobility. We used Stata 16 for analysis.

## RESULTS

### COVID-19 Impact on Emergency Department Visit Volumes and Acuity

In Figure 1, we present trends in overall ED visit volumes by patient age and by ED size. Starting in mid-March (week 11), the 2020/2019 visit ratio declined sharply for both adults (age 18 years and older) and children (<18 years), nadiring at around 0.60 for adults and falling under 0.30 for children by mid-April (week 16) (Panel A). ED visits steadily increased to a 2020/2019 ratio of around 0.87 for adults and 0.62 for children by the end of June (week 27). Adult ED visits plateaued from August through the end of September, while visits for children increased slowly over the summer, before declining again by the end of August (week 35). The 2020/2019 visit ratios followed similar trends by ED size, which combine adult and pediatric visits (Panel B). By mid-April (week 16) visit ratios at facilities of all sizes fell to below 0.50 and rebounded to about 0.80 at FSEDs and small EDs and 0.70 at medium and large EDs by mid to late June (weeks 25-27). However, visits then plateaued and began to decline slightly by the end of September.

**Figure 1.**
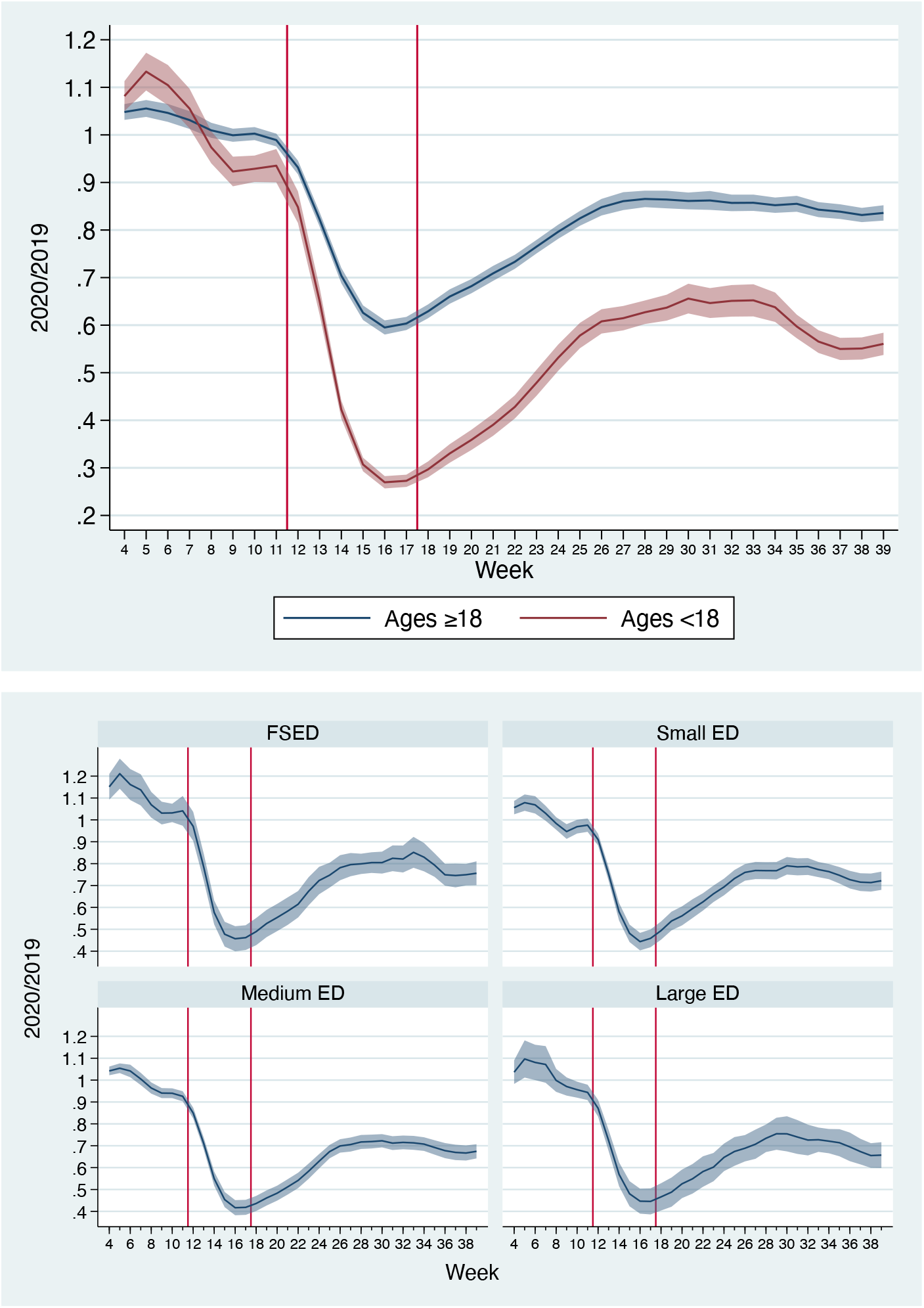
2020/2019 Ratios of visits in 136 EDs (Panel A – top; Panel B – bottom) 2020/2019 ratios from 24 free standing EDs, 40 small EDs (<30,000 visits/y), 56 medium EDs (30,000-59,999 visits/year), and 16 large EDs (>60,000 visits/y). Red vertical lines divide the pre-COVID, early COVID, and COVID time periods. Shaded lines are 95% confidence intervals. ED = emergency department; FSED = freestanding ED

In Figure 2, the impact of COVID-19 on visit acuity (RVUs/visit) is presented. For children, the 2020/2019 ratio of RVUs/visit increases to 1.2 by the end of April and to just under 1.1 for adults before declining to around 1.1 for children and to 1.04 for adults by the end of June (week 27). RVUs/visit then begin to increase slightly, and remain above 1.0, through the end of September. Combining adults and pediatric visits, trends in acuity were similar by ED size (Panel B).

**Figure 2.**
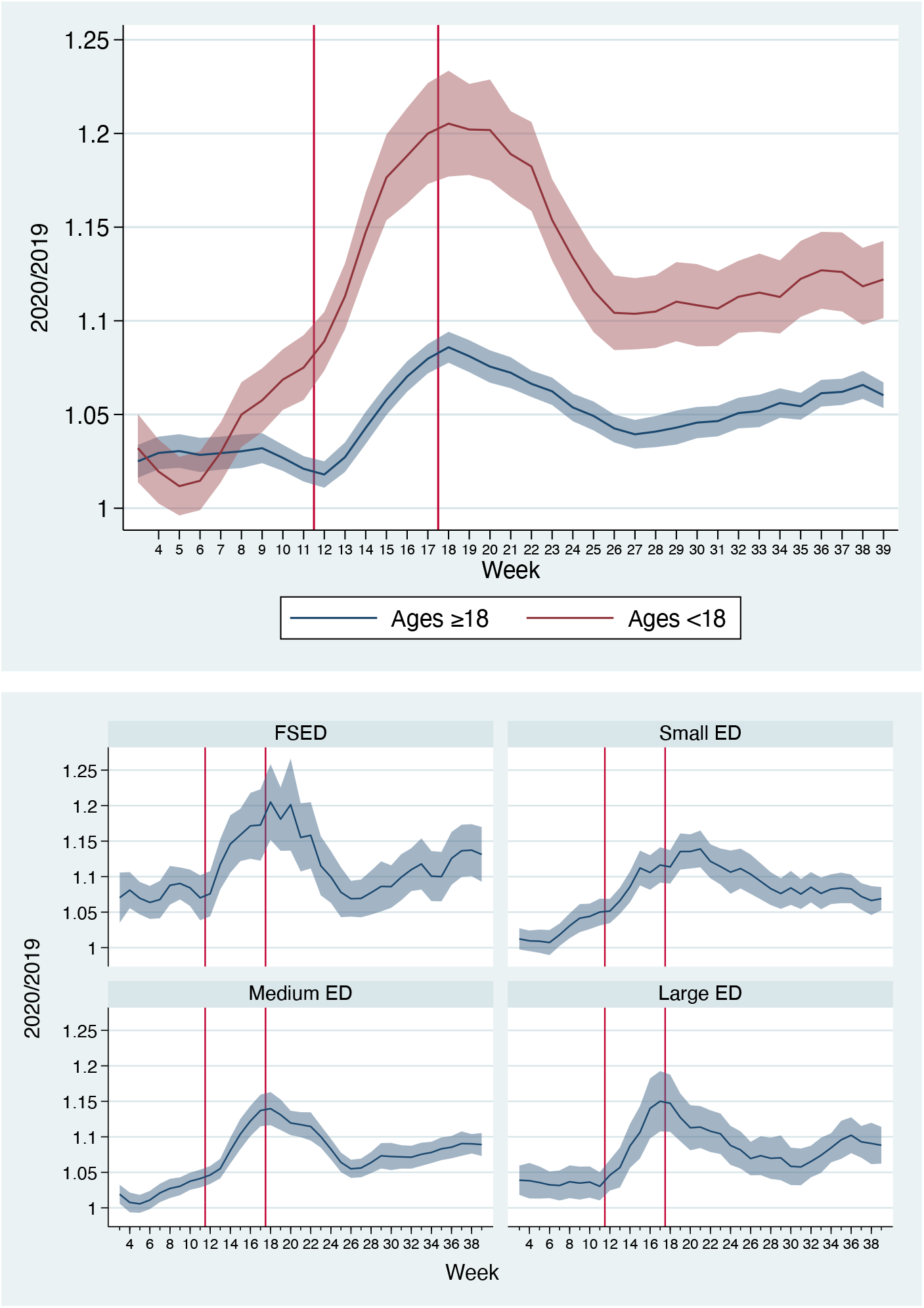
2020/2019 Ratio of RVUs per visit in 136 EDs (Panel A – Top; Panel B – Bottom) 2020/2019 ratios from 24 free standing EDs, 40 small EDs (<30,000 visits/y), 56 medium EDs (30,000-59,999 visits/year), and 16 large EDs (>60,000 visits/y). Red vertical lines divide the pre-COVID, early COVID, and COVID time periods. Shaded lines are 95% confidence intervals. ED = emergency department; FSED = freestanding ED; RVU = relative value unit

### COVID-19 Impact on Emergency Care Expenses and Revenues

In Figure 3, we present revenues and direct expenses by ED size. Direct expenses for FSEDs and small EDs declined slower and less significantly than in medium and large EDs. For these smaller EDs, 2020/2019 expense ratios declined to approximately 0.88 by mid-May, increasing slightly to 0.91 by the end of September. Conversely, 2020/2019 expense ratios for medium and large EDs decreased to 0.58 by mid-April, increasing to 0.87 by the end of September. Revenues for all ED types fell sharply during the early-COVID period and subsequently recovered, but plateaued below 2019 levels. For FSEDs and small EDs, 2020/2019 revenue ratios declined to 0.62 and 0.57 by mid-April (week 16) and increased to 0.93 and 0.88 by the end of September (week 39), respectively. 2020/2019 revenue ratios for medium and large EDs followed a similar trend, nadiring at around 0.58 by mid-April and increasing to 0.87 by the end of September.

**Figure 3.**
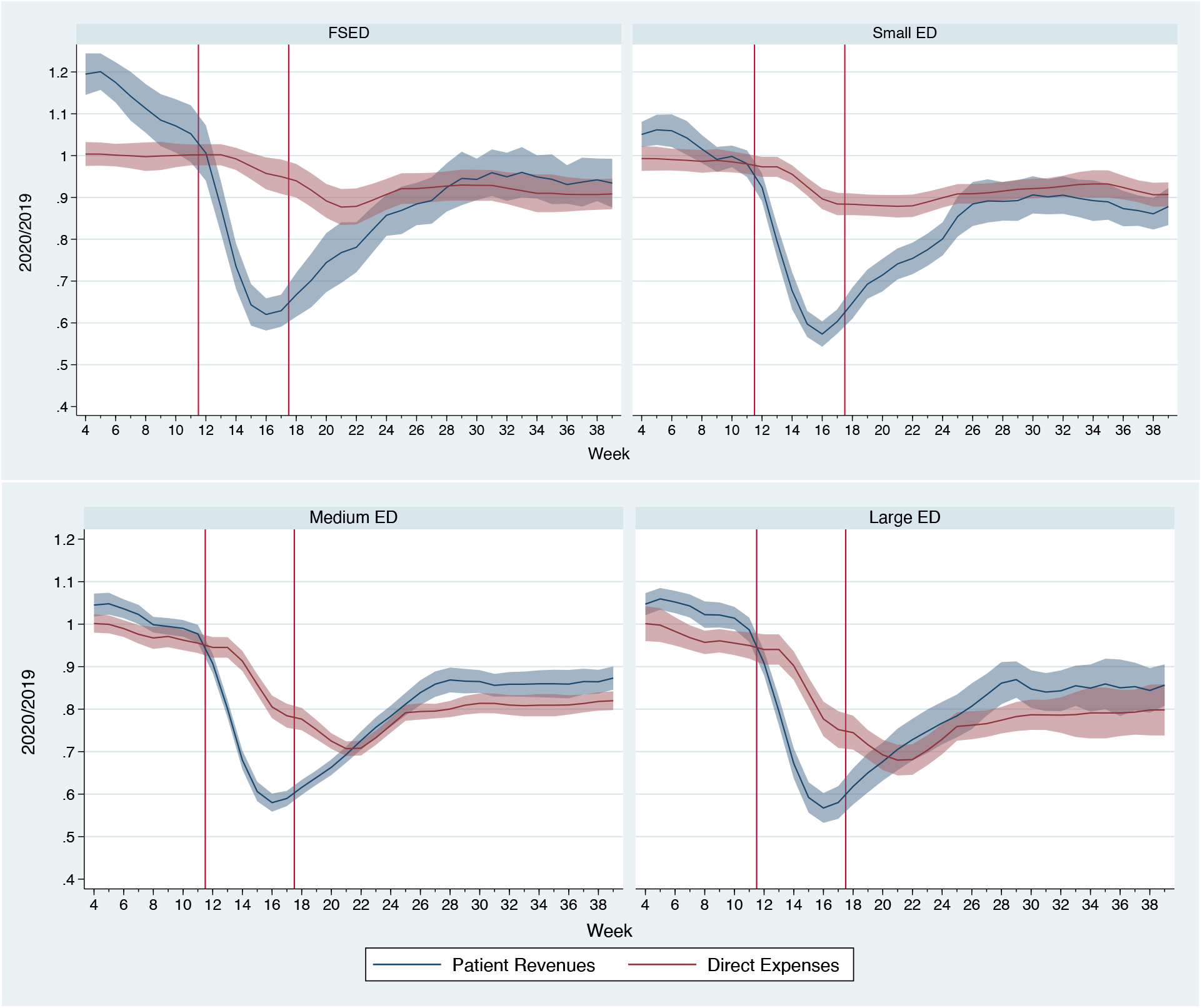
2020/2019 Ratios of patient revenues and direct expenses in 136 EDs. 2020/2018 ratios from 24 free standing EDs, 40 small EDs (<30,000 visits/y), 56 medium EDs (30,000-59,999 visits/year), and 16 large EDs (>60,000 visits/y). Red vertical lines divide the pre-COVID, early COVID, and COVID time periods. Shaded lines are 95% confidence intervals. ED = emergency department; FSED = freestanding ED

### COVID-19 Impact on Clinician Workload

The impact of COVID-19 on hours worked for both physicians and APPs is presented in Figure 4 and in Table 1. As seen in the Figure, the 2020/2019 physician hour ratio at FSEDs and small EDs remained relatively steady at around 1.0, indicating no reduction in physician hours at those sites relative to 2019. Conversely, 2020/2019 APP hour ratios declined significantly at those sites, following below 0.30 at FSEDs and below 0.50 at small EDs, and increasing to around 0.60 by the end of September. At medium and large EDs, however, 2020/2019 physician hour ratios fell under 0.80 at medium EDs and under 0.70 at large EDs by late April/early May (weeks 19-20), before increasing to 0.85 at medium EDs and 0.77 at large EDs by the end of September. APP hours also fell at these larger sites, although not to the extent as seen in the smaller facilities (2020/2019 APP hour ratios nadired at 0.64 and 0.68 at medium and large EDs, respectively).

**Table 1.** Changes to the Emergency Medicine Clinician Workforce During the COVID-19 Pandemic in 136 EDs.

|  | Mean hours<br>worked per<br>week pre-COVID-19 | Mean hours<br>worked per<br>week during<br>COVID-19 | Mean change<br>in hours<br>worked per<br>week | %<br>reduction<br>in hours<br>worked | Total hours<br>reduced during<br>COVID-19<br>(28 weeks) | Total<br>reduction<br>in FTEs |
| --- | --- | --- | --- | --- | --- | --- |
| **Physician workforce** |  |  |  |  |  |  |
| Free Standing EDs | 4,057 | 3,909 | -148 | -3.6 | 4,144 | 5.5 |
| Small EDs | 6,397 | 6,158 | -239 | -3.7 | 6,692 | 8.9 |
| Medium EDs | 14,261 | 11,719 | -2,542 | -17.8 | 71,176 | 94.1 |
| Large EDs | 6,748 | 4,969 | -1,779 | -26.4 | 49,812 | 65.9 |
| **Total** | **31,463** | **26,755** | **-4,708** | **-15.0** | **131,824** | **174.4** |
| **APP workforce** |  |  |  |  |  |  |
| Free Standing EDs | 589 | 302 | -287 | -48.7 | 8,036 | 10.6 |
| Small EDs | 2,187 | 1,418 | -769 | -35.2 | 21,532 | 28.5 |
| Medium EDs | 11,089 | 8,379 | -2,710 | -24.4 | 75,880 | 100.4 |
| Large EDs | 5,738 | 4,286 | -1,452 | -25.3 | 40,656 | 53.8 |
| **Total** | **19,603** | **14,385** | **-5,218** | **-26.6** | **146,104** | **193.3** |

**Note: Clinician hours worked show totals/means across 24 FSEDs, 40 small EDs (<30,000 visits/y), 56 medium EDs (30,000-59,999 visits/year), and 16 large EDs (>60,000 visits/y). Full-time equivalents (FTEs) are defined at 108 hours worked/month (27 hours per week). Pre-COVID-19 consists of first 11 weeks of the year (excluding week 1) through March 14, 2020. COVID-19 period consists of 28 weeks, from March 15 to September 26.**

**Figure 4.**
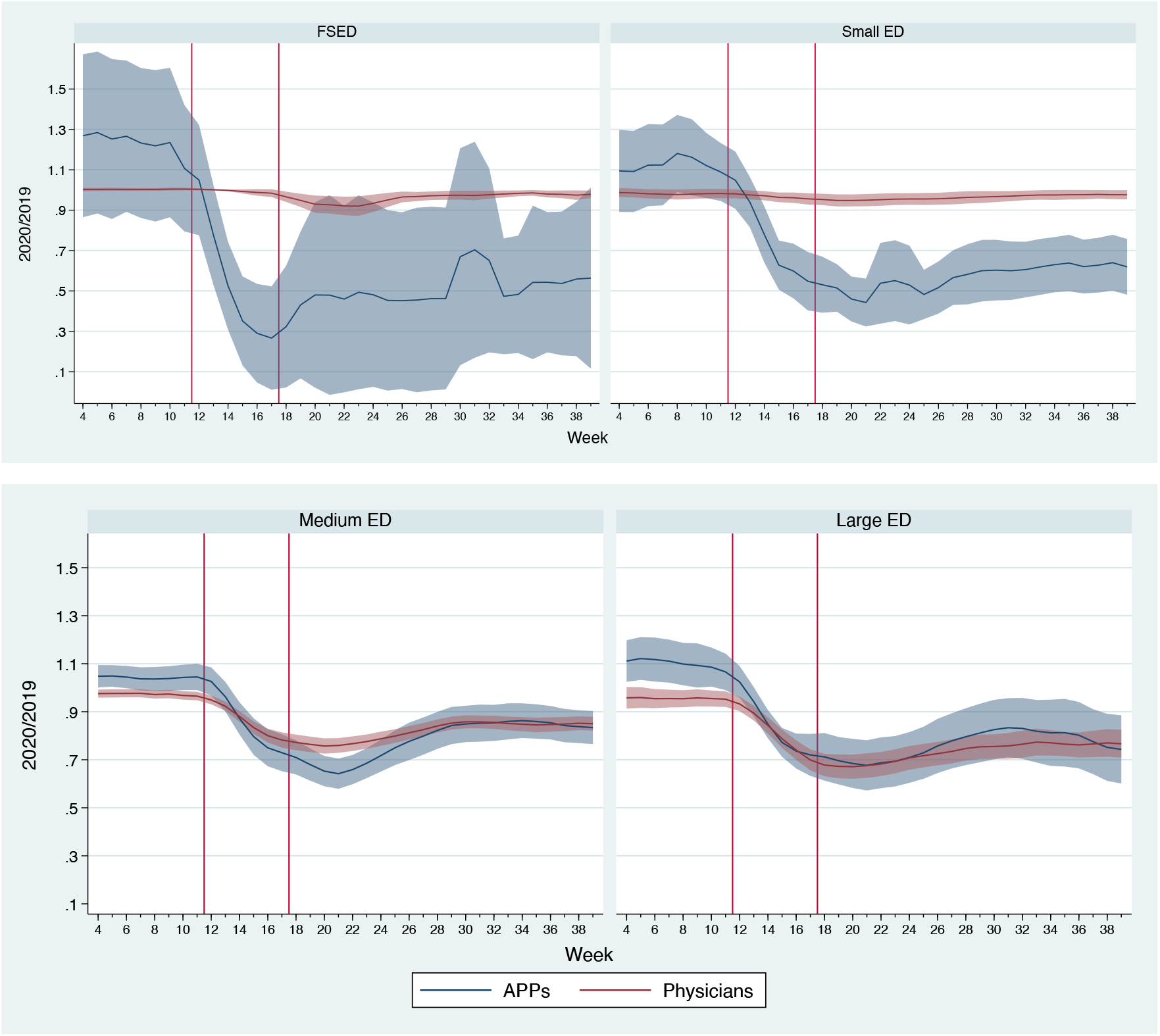
2020/2019 Ratios of clinician hours in 136 EDs. 2020/2018 ratios from 24 free standing EDs, 40 small EDs (<30,000 visits/y), 56 medium EDs (30,000-59,999 visits/year), and 16 large EDs (>60,000 visits/y). Red vertical lines divide the pre-COVID, early COVID, and COVID time periods. Shaded lines are 95% confidence intervals. ED = emergency department; FSED = freestanding ED

In Table 1, we quantify the total impact of the reductions in work hours during COVID-19, compared to the pre-pandemic period in 2020 (Jan 5-March14). Across all sites, physician hours were reduced 15% and APP hours 27% during COVID-19, translating to a loss of 174 physician FTEs and 193 APP FTEs. Physician hours were cut most significantly at large EDs (−26%) and medium EDs (−18%) whereas APP hours were cut most significantly at FSEDs (−49%) and small EDs (−35%).

## LIMITATIONS

There are several limitations to our study. First is the generalizability of our findings outside of the 136 sites, which represent about 2% of U.S. EDs. More broadly, the need to reduce hours to match volume was a widely recognized necessity among ED practice groups during the early pandemic. However, this national group did not reduce hourly payments to clinicians. Therefore, we may overestimate the economic effect in other groups that reduce hourly pay but underestimate the impact on clinicians. Second, we used expected revenue because as of this article’s writing, payments had not yet been collected due to the longer nature of the revenue cycle where collections can sometimes be considerably delayed. While expected revenues tracks closely with actual revenue based on historical experience, it is possible that results may be slightly different after actual revenue is collected. Third, we did not account for any costs of management in this analysis, only clinical revenues from insurance reimbursement and patient payments and direct expenses for salaries (i.e. gross margin). The actual expenses of managing a large group of physicians is considerably higher. Therefore, our results may overestimate the real marginal profitability of EDs, particularly when solely comparing clinical revenues and clinical expenses.

Fourth, we did not account for any stipends provided by hospitals to supplement revenue, which may underestimate total payments to maintain each site. Fifth, these data did not account for the Coronavirus Aid, Relief, and Economic Security Act (CARES) funding that was provided to the national group by the federal government during the pandemic. There was a single large payment to the group from the CARES Act, which only offset an estimated 27% of the revenue impact of the pandemic (applied to weeks 12-48 of 2020). In addition, CARES Act funding provided additional payments for uninsured patients evaluated for COVID-19 or COVID-19 related symptoms which offset the revenue impact by an additional 3%. We also did not account for revenues or costs related to the use of telemedicine which also increased during the pandemic. However, this only represented a small percentage of clinical revenue in this group and did not increase profit margins. Sixth, we did not account for the five EDs that closed temporarily or permanently during the COVID-19 pandemic.

## DISCUSSION

Our data demonstrates the vulnerability of the economics of a healthcare system that solely reimburses emergency clinician services through fee-for-service payments. These effects are driven by the large declines in visits observed for both adult and, more so, pediatric visits early in the pandemic that subsequently increased yet have remained depressed compared to 2019. ED practice organizations, along with the national staffing group, reduced clinical hours to match lower volumes of patients with lower clinical hours. Notably, another relative decline in ED visits occurred in September likely due to the fact that many schools remained closed. In the face of this economic hardship, the average acuity of patients in the ED increased for both adults and children, with the remaining volume of patients being sicker on average, presumably due to lower acuity patients either avoiding ED care or seeking treatment through other pathways (e.g., telemedicine). Higher acuity visits increased clinical revenue

In our data, the economics of small EDs and FSEDs appear to be more vulnerable under the current reimbursement model than medium and large EDs. Even after the early pandemic, these facilities have remained either breakeven (for FSEDs) or unprofitable (for small EDs), before any management fees or stipends. These facilities remain economically stressed because a minimum of single 24/7/365 emergency physician coverage is required in all facilities, despite lower volumes. This brings into question the long-term sustainability of continued staffing of these facilities with 24/7/365 physician coverage, without considerable stipends. To date, a handful (4) FSEDs closed for a short period which were excluded from our study, but later reopened while one FSED has closed permanently. The impact on lower volume EDs is of particular concern when considering that smaller hospital-based EDs commonly serve rural or other under-served communities.

During the pandemic, there has been a 15% decline in the workforce needs for physicians and 26% for APPs to match the volume demands. This translated to a loss of 174 and 193 FTE positions, respectively. Lowered workforce needs translated to a combination of lowering clinician hours (and salaries), and all-cause departures of 58 APPs and 62 physicians from the national group since mid-March. Graduates of residency programs ending in both 2020 and 2021 are experiencing a challenging job market, as a likely result of a generalizable effect of what we observe in our data. These negative economic effects have occurred notably at the same time when the emergency medicine community has served as the frontline during a national public health emergency, increasing risks to their own health through exposure to a novel virus with concurrent limitations in the availability of personal protective equipment. In addition, there is a planned 6% reduction in Medicare reimbursement for emergency physician clinical services starting in 2021 based on the announced physician fee schedule from the Centers for Medicare and Medicaid Services, which will further worsen the economics of ED care.^6^

The economic effects and the staffing responses to the pandemic we observed differed by ED size. Smaller facilities and FSEDs lowered costs primarily through furloughing APPs. For medium and large facilities, there was a combination of physician and APP staffing reductions. A potential effect may be a long-term shift of ED-trained APPs into other facilities or even other fields of medicine, given the ability of APPs to switch fields more easily than physicians. For medium and large EDs, the pandemic caused dramatic declines in payments and 2-3 months where expenses dramatically exceeded payments. However, these facilities have remained more economically viable than small EDs and FSEDs, but at the expense of clinicians’ salaries and some jobs.

Our data have implications for the funding of EDs longer-term post-pandemic. First, we demonstrate the vulnerability of a fee-for-service system in funding facilities that serve the public as the first line of defense during pandemics and as a safety net for the uninsured. Policymakers should consider our data when creating policies about how to ensure EDs are properly staffed for both everyday care as well as public health emergencies like the COVID-19 pandemic, in particular smaller EDs and FSEDs that serve less populous and/or more rural areas. Given the high fixed costs of staffing 24/7/365, reimbursement for capacity and readiness needs to be included in the payment model for emergency care to ensure financial sustainability. Finally, our data suggests a need for a potential shift of payments for emergency clinicians and EDs to alternative payment models. One relevant example is global budgets which are used in Maryland hospitals, which have reduced their susceptibility to the large volume shifts observed during the COVID-19 pandemic.^7^

In conclusion, the COVID-19 pandemic has had a dramatic impact on the economics of EDs and the livelihood and job prospects for emergency physicians and APPs. After a period of expenses exceeding clinical payments, medium and large EDs have been able to recover financially through lower ED staffing with fewer clinical hours for both physicians and APPs. Small and FSEDs have remained unprofitable or break-even bringing into question their long-term viability, particularly if volumes remain depressed for a longer period of time without additional sources of funding beyond the fee-for-service system.

## Data Availability

The data referred to in this study are not publicly available.

---

**Appendix Table.** Facility-Level Means and Ratios, 2019 and 2020.

|  | **Pre-COVID-19** |  | **Early COVID-19** |  | **COVID-19** |  |
| --- | --- | --- | --- | --- | --- | --- |
|  | **January 5 - March 14** |  | **March 15 - April 25** |  | **April 26 - September 26** |  |
|  | **2019** | **2020** | **2019** | **2020** | **2019** | **2020** |
| **Visits per Week** |  |  |  |  |  |  |
| All facilities | 370.8 | 370.3 | 367.5 | 214.2 | 356.4 | 264.6 |
| Facility Sizes |  |  |  |  |  |  |
| Free Standing | 133.3 | 141.8 | 134 | 77.2 | 132.3 | 101.3 |
| ED |  |  |  |  |  |  |
| Small ED | 184.3 | 187.8 | 183.2 | 109.3 | 178.8 | 141.0 |
| Medium ED | 419.4 | 416.6 | 419.6 | 252.5 | 412.2 | 311.4 |
| Large ED | 746.1 | 735.0 | 733.2 | 417.9 | 702.4 | 504.5 |
| Patient Age |  |  |  |  |  |  |
| <18 y | 172.9 | 170.6 | 163.9 | 59.2 | 136.4 | 70.6 |
| ≥18 y | 568.7 | 570.0 | 571.2 | 369.2 | 576.4 | 458.6 |
| **RVUs/Visit** |  |  |  |  |  |  |
| All facilities and ages | 3.37 | 3.48 | 3.42 | 3.8 | 3.46 | 3.77 |
| By facility type |  |  |  |  |  |  |
| Free Standing | 3.05 | 3.26 | 3.14 | 3.61 | 3.18 | 3.54 |
| ED |  |  |  |  |  |  |
| Small ED | 3.41 | 3.48 | 3.46 | 3.77 | 3.48 | 3.78 |
| Medium ED | 3.52 | 3.59 | 3.55 | 3.9 | 3.58 | 3.86 |
| Large ED | 3.49 | 3.60 | 3.53 | 3.92 | 3.6 | 3.9 |
| By patient age |  |  |  |  |  |  |
| <18 y | 2.79 | 2.92 | 2.88 | 3.38 | 2.95 | 3.34 |
| ≥18 y | 3.94 | 4.05 | 3.96 | 4.22 | 3.97 | 4.2 |
|  | **January 5 - March 14** |  | **March 15 - April 25** |  | **April 26 - September 26** |  |
|  | **Revenues** | **Expenses** | **Revenues** | **Expenses** | **Revenues** | **Expenses** |
| **2020/2019 Ratios** |  |  |  |  |  |  |
| All facilities | 1.04 | 0.98 | 0.70 | 0.91 | 0.83 | 0.84 |
| Free Standing EDs | 1.13 | 1.00 | 0.75 | 0.98 | 0.88 | 0.91 |
| Small EDs | 1.02 | 0.98 | 0.70 | 0.93 | 0.84 | 0.91 |
| Medium EDs | 1.01 | 0.97 | 0.69 | 0.88 | 0.81 | 0.79 |
| Large EDs | 1.03 | 0.97 | 0.69 | 0.86 | 0.80 | 0.76 |
|  | **APP Hours** | **Physician Hours** | **APP Hours** | **Physician Hours** | **APP Hours** | **Physician Hours** |
| **2020/2019 Ratios** |  |  |  |  |  |  |
| All facilities | 1.08 | 0.97 | 0.80 | 0.91 | 0.70 | 0.87 |
| Free Standing EDs | 1.23 | 1.00 | 0.54 | 0.99 | 0.51 | 0.96 |
| Small EDs | 1.11 | 0.98 | 0.76 | 0.97 | 0.57 | 0.96 |
| Medium EDs | 1.04 | 0.97 | 0.86 | 0.86 | 0.78 | 0.82 |
| Large EDs | 1.09 | 0.95 | 0.84 | 0.82 | 0.76 | 0.73 |

N = 24 free standing EDs, 40 small EDs (<30,000 visits/y), 56 medium EDs (30,000-59,999 visits/year), and 16 large EDs (>60,000 visits/y).

